# Atherosclerotic fibrous plaques in females are characterized by endothelial-to-mesenchymal transition and linked to smoking

**DOI:** 10.1101/2024.10.01.24314739

**Authors:** Tim R. Sakkers, Eloi Mili, Denitsa Meteva, Marian Wesseling, Daniek Kapteijn, Barend M. Mol, Gert J. de Borst, Dominique P.V. de Kleijn, Sander W. van der Laan, Mete Civelek, Manuel Mayr, Gerard Pasterkamp, Michal Mokry, Ernest Diez Benavente, Hester M. den Ruijter

**Author notes:** **Corresponding author: Hester M. den Ruijter**, PhD Division of Heart & Lungs, Department of Cardiology, Laboratory of Experimental Cardiology. These authors contributed equally (Tim R. Sakkers, Ernest Diez Benavente, Hester M. den Ruijter).

## Abstract

**Background:** Sex and plaque histology are intertwined, with fibrous atherosclerotic plaques being more prevalent in women and pointing to general smooth muscle cell plasticity and estrogen signaling. Plaque erosion, a significant contributor to acute coronary syndromes (ACSs), is linked to fibrous plaques and is more prevalent in women as compared to men. We hypothesize that the molecular drivers of histologically determined fibrous plaques differ between men and women.

**Methods:** Human end-stage atherosclerotic plaques were isolated from consecutive patients who underwent carotid endarterectomy and were included in the Athero-Express biobank. Fibrous plaques from both female and male patients were histologically assessed and further processed to obtain protein, bulk RNA, single-cell RNA and DNA methylation data. We leveraged sex-differential expression and deconvolution analyses to uncover sex-biased molecular mechanisms and cellular dynamics which were experimentally validated using an EndMT in vitro model.

**Results:** Out of 1,889 atherosclerotic plaques (1,309 male and 580 female), fibrous lesions were observed in 50% of female (n=290) and 31% of male patients (n=416). Compared to patients with atheromatous plaques (n=494), women with fibrous plaques exhibited a higher prevalence of smoking (41% vs. 33%), while men with fibrous plaques presented more often with diabetes (29% vs. 20%). Transcriptional and proteomic phenotyping highlighted more immune-dependent and inflammatory processes in male fibrous plaques. Genes and proteins with higher abundance in female fibrous plaques pointed to endothelial-to-mesenchymal transition (EndMT) and extracellular matrix remodelling. Using single-cell RNA sequencing, we identified a dominant role of endothelial and smooth muscle cells in female plaques, and more macrophages in males. Finally, at the cellular level, we show that sex - specific, smoking-mediated promoter methylation changes may explain these differences.

**Conclusions:** Patients with end-stage fibrous atherosclerotic plaques have a distinct clinical profile, with men more often having diabetes and women more often smoking. This clinical profile associates with sex differences that point to different cellular and molecular compositions of fibrous plaques. These mechanisms might be candidate pathways to understand plaque erosion from a molecular point of view and may provide promising targets for atherosclerosis therapies, as they account for the sex-specific differences in plaque phenotype.

**Graphical abstract:** 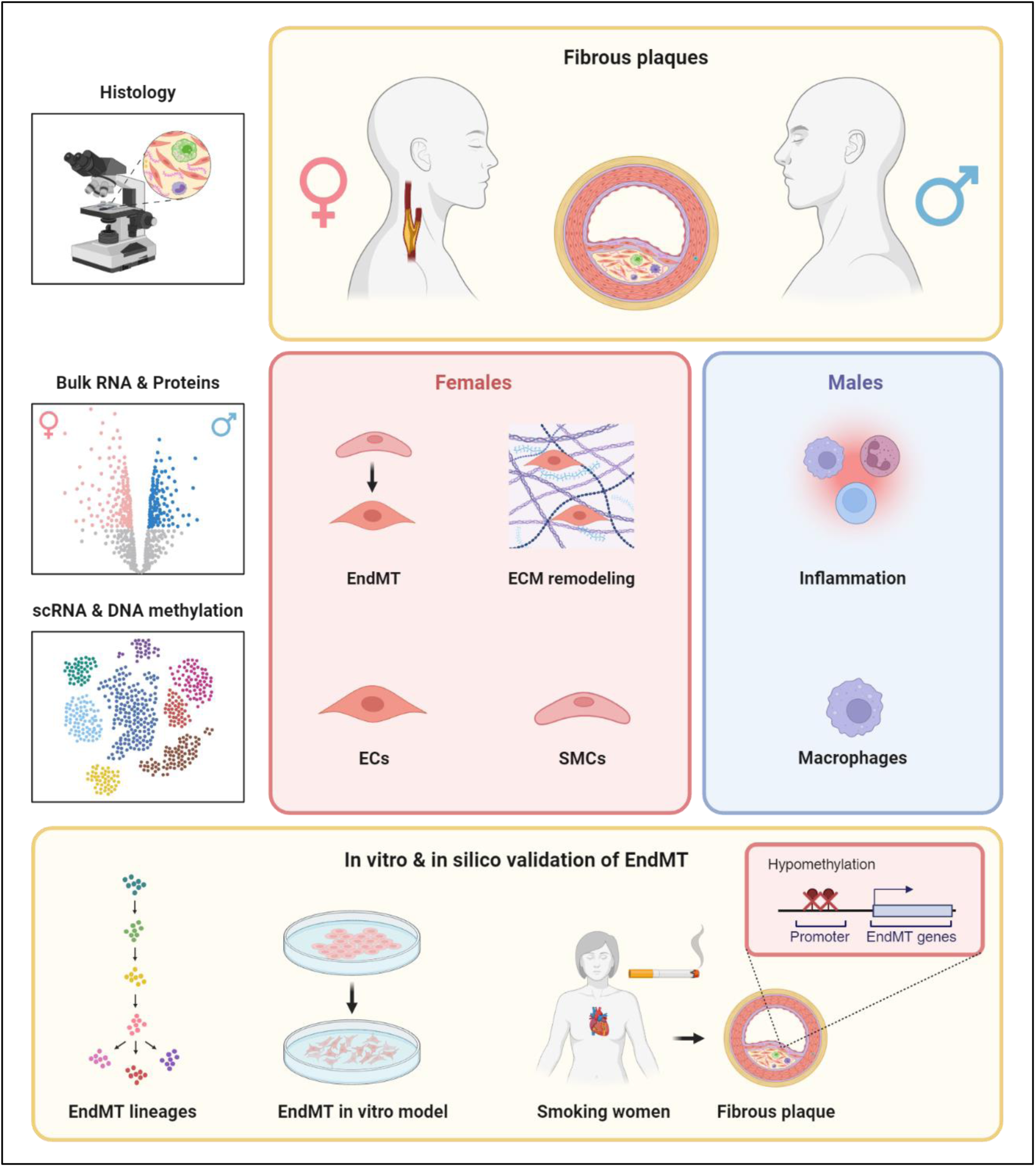

## Introduction

Plaque erosion significantly contributes to the incidence of acute coronary syndromes (ACSs) with a pronounced prevalence in women (51% vs. 28% in men), especially among young women (<50 years: 77% vs. 35% in men)^1^. Plaque rupture is more common in men (69% vs. 37% in women) and is associated with elevated blood cholesterol ^1^. In the current era of widely available lipid-lowering therapies, the proportion of ACSs attributable to plaque erosion may be on the rise in both women and men ^2,3^. Plaque erosion is often linked to non-ST segment myocardial infarction (NSTEMI), in contrast to plaque rupture, which is more frequently the cause of ST segment myocardial infarction (STEMI) ^2^.

The mechanism of plaque erosion is thought to be endothelial dysfunction on top of fibrous plaques, which can trigger acute thrombosis ^4–7^. Women presenting with severe atherosclerotic disease more often present with fibrous plaques, while symptomatic fibrous plaques are also found in men, particularly young men ^8,9^. Fibrous plaques are considered “stable” plaques characterized by high collagen and smooth muscle cell (SMC) content. Yet, fibrous plaques can become symptomatic by mechanisms that promote plaque erosion.

From a clinical perspective, diabetes is recognized as a pivotal risk for atherosclerosis, primarily linked to inflammation-driven unstable plaques prone to rupture ^10^. In contrast, smoking is another well-established risk factor for atherosclerosis, and has been linked to plaque erosion in women ^1^. Notably, prolonged smoking leads to worse cardiovascular outcomes in women compared to men, but those mechanisms are yet to be fully understood ^11,12^. One potential mechanism could involve epigenetic regulation, as it has been shown that plaque methylation is strongly influenced by smoking ^13^. Despite the clinical importance of investigating the mechanism of plaque erosion, symptomatic fibrous plaques are currently understudied.

As women more often present with fibrous atherosclerotic plaques, sex-stratified analyses of atherosclerosis have shed light on potential mechanisms of such plaques. We and others have shown that female atherosclerosis is characterized by gene regulatory networks (GRNs) distinct from men. These GRNs are active in both endothelial cell (EC) and smooth muscle cell (SMCs) plasticity, such as endothelial-to-mesenchymal transition (EndMT) ^14,15^. EndMT is an important process in atherosclerosis where endothelial cells acquire mesenchymal characteristics, leading to increased cellular migration, invasion, and ECM production ^16,17^. Furthermore, these GRNs are enriched for estrogen receptor signaling, suggesting a role for sex hormones in the observed differences between female and male plaques ^14,15^. Sex and plaque phenotype are intimately linked and, therefore, difficult to disentangle. However, the specific differences between sexes within the same plaque phenotype and the impact of associated risk factors remain unclear. Given the increasing prevalence of NSTEMI and fibrous atherosclerotic plaques over the last decades^2,3^, sex-specific studies may provide important novel insights into this critical part of plaque pathology. We hypothesized that sex differences exist within these fibrous lesions, with female fibrous plaques mainly driven by fibrotic processes, while male fibrous plaques are more influenced by immune-related mechanisms.

Therefore, we obtained human end-stage atherosclerotic plaques and specifically examined fibrous lesions classified by their histological features. We studied risk factor profiles of these patients and explored sex differences within plaques at protein, gene, and DNA methylation levels. We provide evidence that female fibrous plaques are characterized by SMC-driven ECM remodelling and EndMT, which is likely linked to smoking-induced changes in epigenetics. Male fibrous plaques, on the other hand, point to inflammation driven by macrophages and diabetes as an important risk factor.

## Methods

### Patient samples

The Athero-Express Biobank (AE) is an ongoing longitudinal biobank since 2002 aimed at investigating atherosclerotic plaques in patients undergoing arterial endarterectomy. Clinical data is obtained through baseline blood withdrawal and extensive questionnaires filled in by participants that are verified against medical records. We analyzed patients who underwent carotid endarterectomy (CEA) and had available protein, gene, and DNA methylation plaque data. All aspects of sample collection, plaque histology, DNA/RNA extraction and sequencing were previously described ^18–20^. The performed study was conducted in accordance with the Declaration of Helsinki^21^, and informed consent was provided by all study participants after approval for this study by the medical ethical committees of the two Dutch tertiary referral centers (UMC Utrecht & St. Antonius Hospital). A composite cardiovascular end point was used for the baseline table. This consisted of (sudden) cardiovascular death, stroke, myocardial infarction, coronary intervention (coronary artery bypass grafting or percutaneous coronary intervention), peripheral reintervention or leg amputation.

### Plaque histology

The atherosclerotic plaque was immediately processed following surgical removal. As previously documented ^14,18,22^, (immune-)histochemical staining was routinely performed on the segment with the most significant plaque burden (culprit lesion) for identification of macrophages (CD68), calcification (hematoxylin-eosin), SMCs (alpha actin), collagen (picro sirius red), plaque hemorrhage (hematoxylin-eosin, Elastin von Gieson staining), vessel density (CD34), and fat (picrosirius red, hematoxylin-eosin). Plaques underwent a semiquantitative assessment, and their classification into fibrous, fibroatheromatous, or atheromatous plaques was carried out in accordance with previously described scoring protocol ^18^. The consistency between and among observers was examined previously and showed good concordance (κ=0.6–0.9) ^23^. The culprit lesion was used for histology, while the plaque segments closest to the culprit lesion were used for sequencing. The plaque vulnerability index is a combined score (ranging from 1 to 6) of individual histological features (SMC content, collagen content, lipid content, macrophage content and intraplaque hemorrhage) ^14^. To identify plaque phenotype-specific properties, we focused our analysis on fibrous (Fig. 1) and atheromatous plaques (Supplemental Fig. 1)(excluding fibro-atheromatous plaques), totalling 290 female and 416 male fibrous plaques and 91 female and 403 male atheromatous plaques.

**Figure 1.**
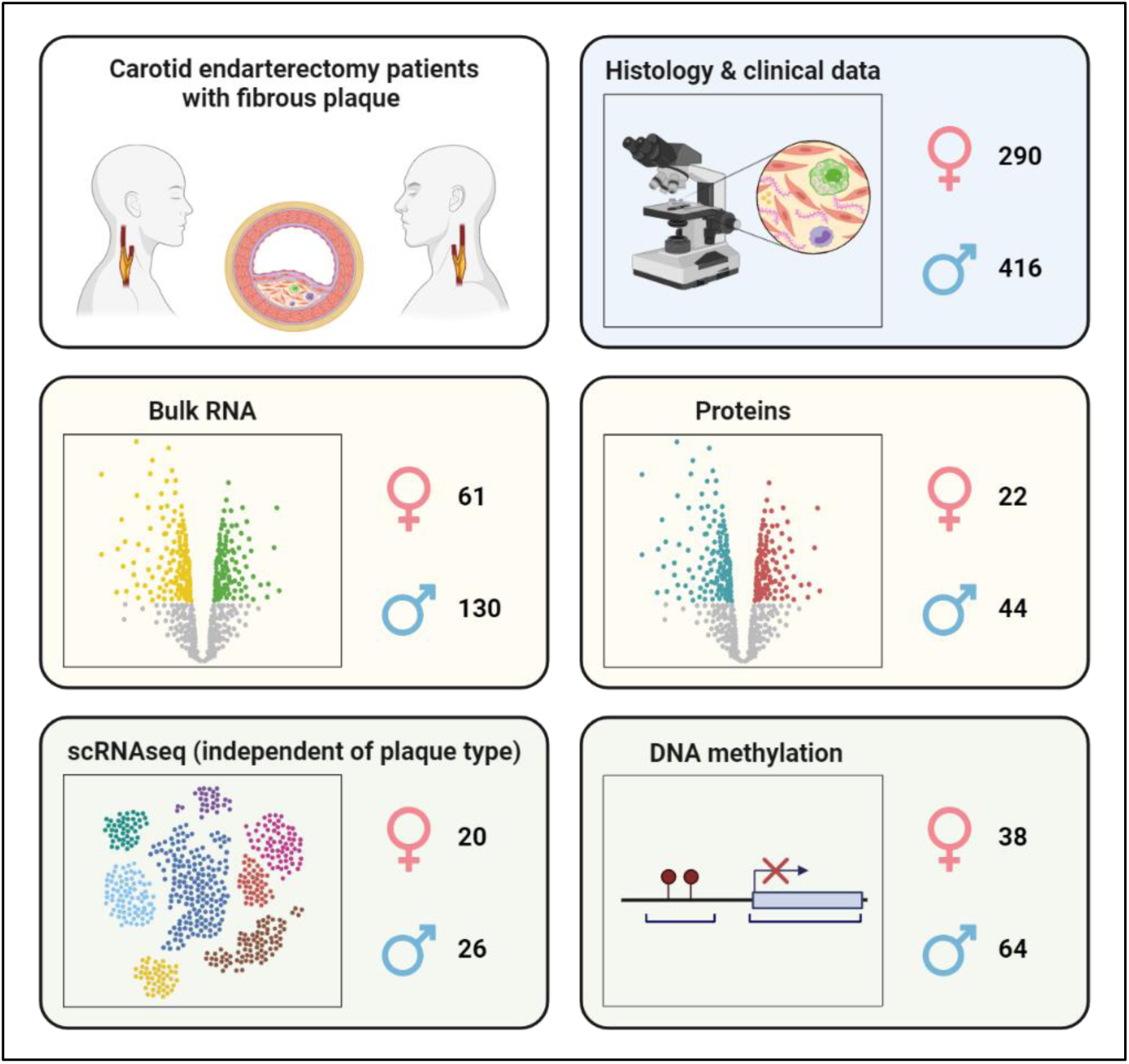
Overview of the patient population with fibrous plaque, including clinical, histology, bulk RNA, protein, scRNAseq and DNA methylation data.

Plaque erosion is defined as the degradation or loss of the endothelial layer covering an atherosclerotic plaque, which can lead to the exposure of the underlying tissue and result in acute thrombus formation. Given its association with fibrous plaques, which are more prone to erosion, these plaques are the primary focus of our study ^4–7^.

### Statistical analysis

All analyses were performed using R (version 3.6.2) and its IDE Rstudio (version 1.2 and later). Baseline characteristics of the study population were summarized using the tableone package. For categorical variables, a chi-square test was performed. For normally distributed continuous variables, a two-tailed unpaired Student t test was used, while for non-normally distributed continuous variables, this was a two-tailed unpaired Mann-Whitney U test. The level of significance was set at p < 0.05 for all tests. Some cardiovascular risk factors were not always documented in the electronic health records and contained missing data, including total cholesterol, LDL, HDL, triglycerides, and CRP (Supplemental Table 1 & 3). Analyses were performed on the complete records for these variables.

### Bulk RNA sequencing data

Out of all plaques (n=1,889), 191 plaques (female: 61, male: 130) were used for RNA-sequencing to study sex differences in fibrous plaques at the gene expression level. For the atheromatous plaque comparison, we selected 187 plaques (female: 29, male: 158). Isolation, library preparation, sequencing, and preprocessing of plaque RNA were previously documented ^14,24^. In short, total RNA was extracted from plaque segments using ceramic beads and a tissue homogenizer with use of TriPure. Library preparation for RNA sequencing involved reverse transcription, incorporating a primer mix and unique barcodes for each sample, facilitating pooling of cDNA samples. The cDNA underwent in vitro transcription, primer removal, fragmentation, and purification. RNA quality was assessed using a Bioanalyzer, and cDNA libraries were constructed for sequencing on the Illumina NextSeq500 platform with paired-end reads. Post-sequencing, data were processed for mapping and analysis, employing the Burrows-Wheel Aligner and custom scripts for generating count matrices from sequence reads.

Further analyses were performed using R-3.6.2 and its IDE Rstudio version 1.2 and later. Genes were annotated with Ensembl ID’s and differential gene expression analysis was performed using the DESeq2 R-package ^25^. Including sex chromosomes in our analysis revealed them as the most differentially expressed, accounting for 88% of the top 100 differentially expressed genes (Supplemental Fig. 2A). These genes were removed from subsequent analyses. We selected genes that were differentially expressed (p<0.05, logFC>0.3, baseMean>10) between female and male fibrous plaques (supplemental Fig. 2B) and female and male atheromatous plaques (supplemental Fig. 2C). Differentially expressed genes were enriched using clusterProfiler ^26^.

### Protein data

Untargeted proteomics was conducted on 128 plaques to study sex differences in fibrous (female: 22, male: 44) and atheromatous (female: 9, male: 53) plaques at the protein level. Samples underwent a 2-step protein extraction process, initially with a NaCl buffer to isolate loosely bound proteins, followed by a guanidine hydrochloride (GuHCl) buffer to solubilize mature ECM proteins, with both fractions stored at -80°C. GuHCL extracts were quantified using a Pierce BCA protein assay, and 20µg of protein from each sample was precipitated with ethanol for deglycosylation. A two-step deglycosylation process was employed of the GuHCL extracts to remove glycosaminoglycans and other sugar monomers, preparing the samples for in-solution digestion with trypsin. The digested samples were then purified using a C18 cartridge system and analyzed by LC-MS using an untargeted proteomics approach on a nano-flow LC system. Spectra were collected with an Orbitrap mass analyzer, and data-dependent MS2 scan was performed for the top 15 ions. Data analysis involved a database search against the human UniProtKB/Swiss-Prot database with specific modifications and enzyme settings. Protein identification yielded 2,148 proteins, with ECM and related proteins categorized using Matrisome DB and further in-house selection. Protein abundances were filtered, normalized, and scaled, with missing values imputed using the KNN-Impute method, resulting in a final dataset of 1,499 proteins for further analysis ^27^. Differential abundance analysis was performed using Limma ^28^. We selected proteins that were differentially abundant between female and male fibrous plaques (p<0.05, logFC>0.3, AveExpr>4). Differentially abundant proteins were enriched using clusterProfiler ^26^.

### Single-cell RNA sequencing data

Single-cell RNA sequencing (scRNAseq) was performed on 46 plaques (female: 20, male: 26) to study cell-type-specific expression patterns of differentially expressed genes. The process of preparing, sorting, and sequencing cells from plaques was described in detail previously ^29,30^. scRNAseq data was processed in R-3.6.2 using the Seurat-3.2.2 R-package^31^. Mitochondrial genes and doublets were filtered out from the data, setting thresholds for unique and total reads per cell. Batch effects were corrected using SCTransform. Clustering was performed with 20 principal components, validated through a JackStraw analysis to ensure significant feature distribution. A clustering resolution of 0.8 was chosen to reflect meaningful biological groupings without overfitting. Sensitivity analysis showed minimal impact of clustering parameters on cell-type identification. Optimal clustering parameters were finalized after multiple iterations. Cell populations were identified by analyzing gene expression in individual clusters. Subpopulations of smooth muscle cells (SMCs) and endothelial cells (ECs) were identified by further clustering using 10 principal components at a higher resolution, and distinct identities were assigned based on differential gene expression analyzed via enrichR-3.0 ^32^. The module score for differentially expressed genes was calculated using the addModuleScore function in Seurat ^31^, providing an expression proxy for gene sets. Genes that were not represented in the scRNAseq were removed from this calculation.

### DNA methylation data

DNA methylation data was obtained from 102 fibrous plaques (female: 38, male: 64) to explore cell-type deconvolution and the potential regulation of differentially expressed genes related to EndMT. For the atheromatous plaque deconvolution, we selected 117 plaques (female: 22, male: 95). DNA isolation, preparation, and methylation conversion were previously described ^13,33^. In short, the purity and concentration of DNA were evaluated using the Nanodrop 1000 system (Thermo Scientific). DNA samples were standardized to a concentration of 600 ng/ml, distributed randomly across 96-well plates, and then subjected to bisulfite conversion through a cycling protocol using the EZ-96 DNA Methylation Kit (Zymo Research). Subsequently, DNA methylation was measured on the Infinium HumanMethylation450 Beadchip Array at the Human Genotyping Facility of the Erasmus Medical Center. Quality control, normalization, and filtering of the methylation data were performed following an established cross-package Bioconductor workflow, using the Limma and Minfi packages ^34^. CpG filtering was performed on probes located on sex chromosomes, failed in one or more samples, contained SNPs or mapped to multiple places in the genome.

### Cell-type deconvolution based on DNA methylation

To build our human methylation atlas, we adopted a previously published approach^35^, using methylation data from 39 cell types sorted from 205 healthy tissue samples. In summary, this process involved selecting the top 100 most specific hyper-and hypomethylated CpGs for a selection of cell types (n=8) expected to be present in atherosclerotic plaques: endothelial cells, smooth muscle cells, erythrocyte progenitors, monocytes/macrophages, NK cells, T cells, and B cells. We calculated the proportion of each CpG relative to the overall methylation pattern across these cell types. For each cell type, we then selected the top 100 hypermethylated CpGs with the highest contributions, considering their relative methylation levels. To identify the top 100 hypomethylated CpGs, we used the reversed methylation values (1-beta value). To determine the relative contribution of each cell type (%) to the atherosclerotic plaque, we performed deconvolution using non-negative least squares from the nnls package in R.

### In vitro EndMT model

The stimulation of ECs with TGFβ and TNFα has been widely documented as a model for EndMT ^36,37^. Human coronary artery endothelial cells (HCAECs, Promocell, CAT#C-12221) from three donors (two female and one male) were cultured until 80% confluency and seeded in a 6-well plate with a concentration of 260.000 cells per well. After a 24-hour incubation at 37°C in 5% CO2, cells were treated with human TGFβ alone and a combination of TGFβ with TNFα (10 ng/mL each) in endothelial basal medium MV supplemented with 0.5% fetal bovine serum. Media and stimuli were refreshed after 48 hours. RNA was isolated before stimulation (0 hours) and after 72 hours of stimulation, and cell morphology was monitored throughout via microscopy. Details on staining, qPCR, and RNA analysis can be found elsewhere ^33,38^. Genes were annotated with Ensembl ID’s and differential gene expression analysis was performed using the DESeq2 R-package ^25^.

## Results

### Female and male patients with fibrous plaques have a distinct risk factor profile

For this study, we selected all consecutive patients included in the Athero-Express carotid endarterectomy biobank for which histopathological slides (Fig. 2A) were available between 2002 and 2018 (n=1.889, 31% women). Histopathological assessment revealed 50% of female and 32% of male plaques as being fibrous, and 16% and 31% as being atheromatous, respectively (Fig. 2B). We analysed 290 women and 416 men with fibrous plaques, both with an average age of 68 years (Table 1). Additionally, we compared this cohort to 91 women and 403 men with atheromatous plaques with a mean age of 72 and 70 years old, respectively (Supplemental Table 2). Plaque vulnerability index, which is a combined score of individual histological features (SMC content, collagen content, lipid content, macrophage content, and intraplaque hemorrhage)^14^, did not show differences between men and women (Table 1, Supplemental Table 2). Most women with fibrous plaques presented with a transient ischemic attack (42%) or stroke (23%). These numbers were similar to those in men (41% and 22%, Table 1). Women presenting with fibrous plaques exhibited a higher prevalence of smoking (41% vs. 33%; p=0.039; Fig. 2C), while men with fibrous plaques presented more often with diabetes (29% vs. 20%; p=0.012; Fig. 2D). These differences in risk factor prevalence were specific to patients with fibrous plaques and were not observed in patients with atheromatous plaques (Supplemental Table 2). In addition, higher plasma levels of total cholesterol (5.3 vs. 4.6 mmol/L; p<0.001), LDL (2.9 vs. 2.5 mmol/L; p=0.004), HDL (1.3 vs. 1.1 mmol/L; p<0.001) and C-reactive protein (CRP, 2.0 vs. 1.5 mg/L; p=0.028) were found in women with fibrous plaques. Men presenting with fibrous plaques included a higher proportion with a history of CAD (35% vs. 25%; p=0.004) and had higher plasma levels of eGFR (78.9 vs. 69.0 mL/min; p<0.001). Moreover, significant differences in contralateral stenosis were found between sexes, with men exhibiting more contralateral stenosis. Among patients with atheromatous plaques, HDL, eGFR, and history of CAD were also significantly different between the sexes and similar in direction.

**Figure 2.**
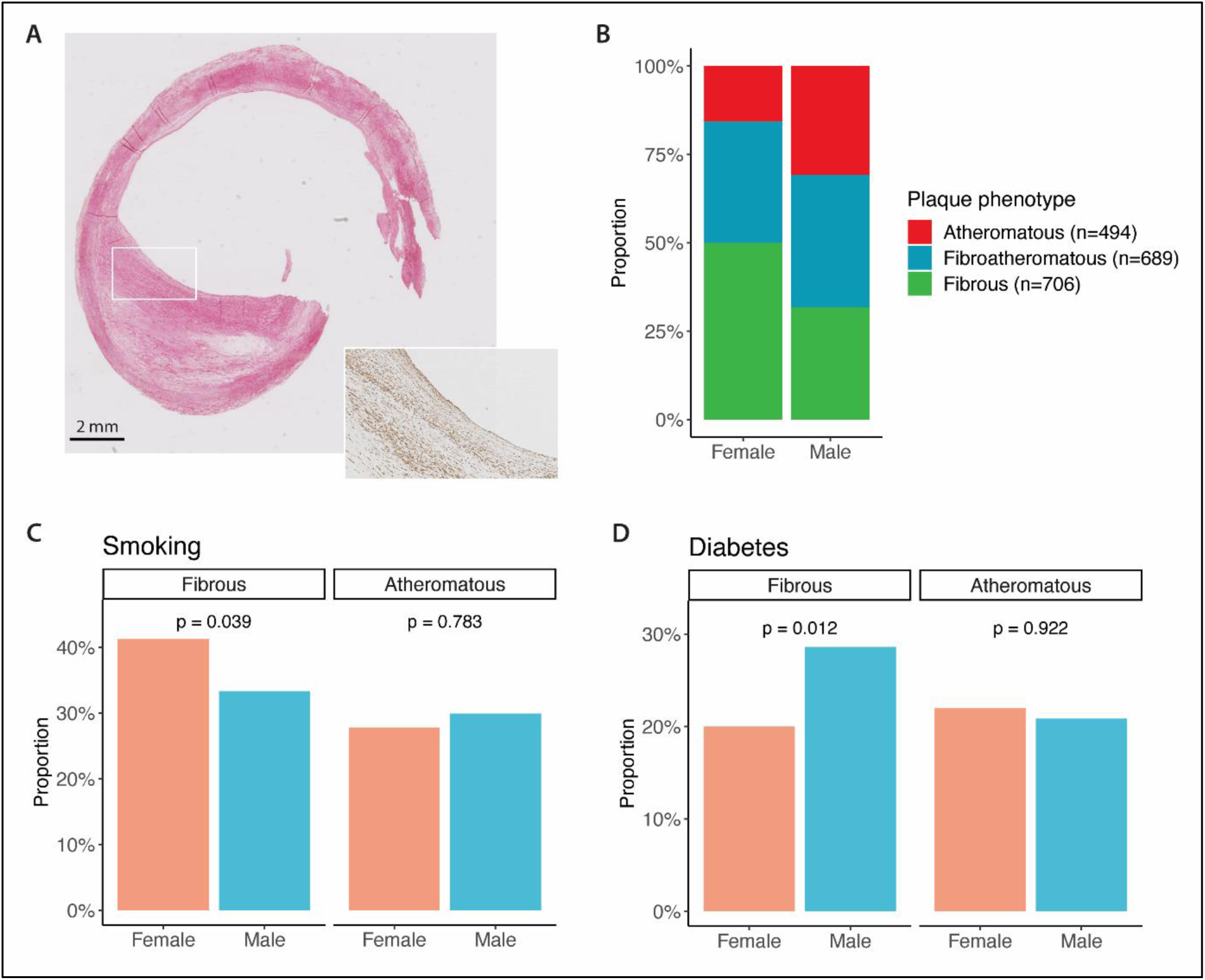
Fibrous plaque with thick fibrous cap containing major staining for collagen (Picrosirius red staining, left) and smooth muscle cells (α-SMA staining, right) (**A**). Distribution of plaque phenotypes in women and men who underwent carotid endarterectomy (**B**). The prevalence of smoking in women and men with fibrous plaques (left panel **C**) versus atheromatous plaques (right panel **C**). The prevalence of diabetes in women and men with fibrous plaques (left panel **D**) versus atheromatous plaques (right panel **D**).

**Table 1:** Clinical characteristics and risk factor profiles of 706 atherosclerosis patients with fibrous plaques. For normally distributed continuous variables, the mean and the SD (in parenthesis) are presented. For non-normally distributed continuous variables, the median and the IQR (in square brackets) are presented. For categorical variables, the counts and percentages (in parenthesis) are presented. Bold P-values are significant differences. BP, blood pressure; LDL, low-density lipoprotein; HDL, high-density lipoprotein; CRP, C-reactive protein; BMI, body mass index; eGFR, estimated glomerular filtration rate; TIA, transient ischemic attack; CAD, coronary artery disease.

| Characteristic | Women (n=290) | Men (n=416) | P-value |
| --- | --- | --- | --- |
| Age (years; mean (SD)) | 68 (9.9) | 68 (9.2) | 0.895 |
| Plaque vulnerability index (mean (SD)) | 2.3 (1.0) | 2.4 (1.0) | 0.326 |
| Systolic BP (mmHg; mean (SD)) | 151 (25.9) | 153 (25.2) | 0.410 |
| Diastolic BP (mmHg; mean (SD)) | 80 (13.1) | 81 (13.9) | 0.309 |
| Hypertension (%) | 214 (75.6) | 292 (71.7) | 0.296 |
| Diabetes (%) | 58 (20.0) | 119 (28.6) | **0.012** |
| Total cholesterol (mmol/L; median [IQR]) | 5.3 [4.3, 6.1] | 4.6 [3.7, 5.6] | **<0.001** |
| LDL (mmol/L; median [IQR]) | 2.9 [2.2, 3.7] | 2.5 [1.9, 3.2] | **0.004** |
| HDL (mmol/L; median [IQR]) | 1.3 [1.1, 1.6] | 1.1 [0.9, 1.4] | **<0.001** |
| Triglycerides (mmol/L; median [IQR]) | 1.4 [1.1, 2.3] | 1.6 [1.1, 2.2] | 0.676 |
| CRP (mg/L; median [IQR]) | 2.0 [1.0, 4.0] | 1.5 [0.7, 3.6] | **0.028** |
| BMI (kg/m<sup>2</sup>; median [IQR]) | 25.9 [23.4, 28.7] | 25.9 [24.0, 28.5] | 0.415 |
| eGFR (mL/min per 1.73 m<sup>2</sup>; mean (SD)) | 69 (24.1) | 79 (25.6) | **<0.001** |
| Smoking (%) | 118 (41.3) | 138 (33.3) | **0.039** |
| Clinical presentation (%) |  |  | 0.093 |
| Asymptomatic | 38 (13.1) | 83 (20.0) |  |
| Ocular | 63 (21.7) | 73 (17.5) |  |
| TIA | 122 (42.1) | 169 (40.6) |  |
| Stroke | 67 (23.1) | 91 (21.9) |  |
| History of CAD (%) | 72 (24.8) | 147 (35.3) | **0.004** |
| Antihypertensive drugs (%) | 213 (73.7) | 324 (78.1) | 0.211 |
| Statins (%) | 232 (80.3) | 330 (79.5) | 0.880 |
| Antiplatelet drugs (%) | 259 (89.6) | 358 (86.5) | 0.256 |
| Anticoagulant drugs (%) | 34 (11.8) | 59 (14.2) | 0.405 |
| Ipsilateral stenosis (%) |  |  | 0.154 |
| 0-50% | 0 (0.0) | 1 (0.2) |  |
| 50-70% | 18 (6.4) | 41 (10.1) |  |
| 70-99% | 265 (93.6) | 363 (89.6) |  |
| Contralateral stenosis (%) |  |  | **0.015** |
| 0-50% | 144 (54.8) | 175 (47.3) |  |
| 50-70% | 36 (13.7) | 54 (14.6) |  |
| 70-99% | 53 (20.2) | 64 (17.3) |  |
| 100% | 30 (11.4) | 77 (20.8) |  |

### Sex differences in gene expression between fibrous plaques point to EndMT and ECM in female plaques and inflammation in male plaques

Fibrous plaques, as defined histologically, may harbour distinct molecular mechanisms beyond their histological characteristics. Therefore, we recently investigated the transcriptomic landscape of atherosclerotic plaques and identified five plaque transcriptomic clusters, including the fibro-collagenous, intermediate, lipomatous, fibro-inflammatory, and fibro-cellular plaque phenotype^24^. We assigned a cluster to each of the plaques within the study based on overall gene expression patterns. In women, 64% of the fibrous plaques matched a fibrous-like phenotype (fibro-collagenous and fibro-cellular), and 11% overlapped with an atheromatous-like phenotype (lipomatous and fibro-inflammatory), while in men, this was 50% and 23%, respectively (Fig. 3A). A similar trend towards inflammatory clusters in men was observed when comparing atheromatous plaques of men and women (Fig. 3A).

**Figure 3.**
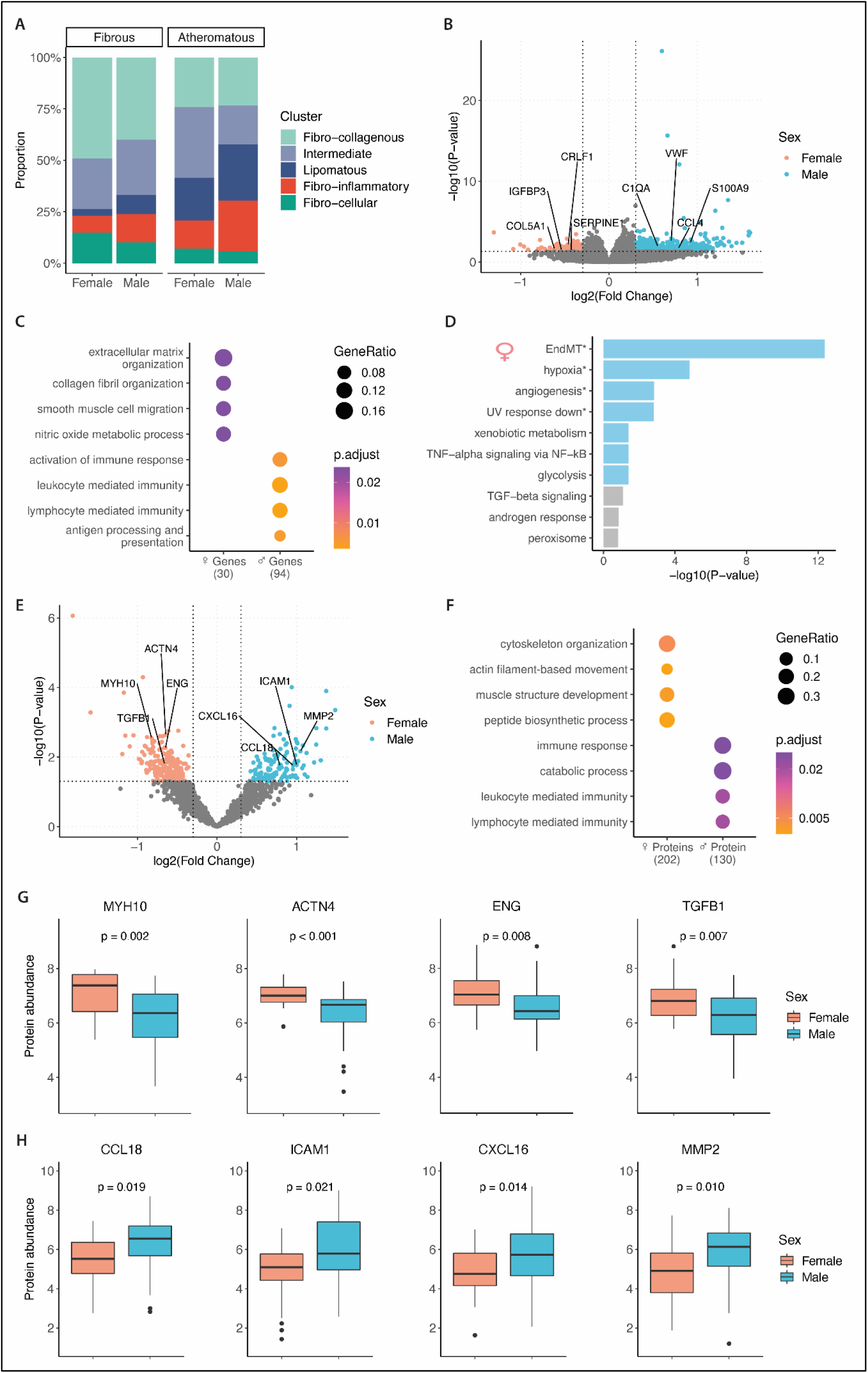
Assignment of atherosclerotic plaques to transcriptomic-based clusters based on gene expression patterns (**A**). A volcano plot is shown for the differential gene expression analysis between fibrous plaques from female and male patients (**B**). GO enrichment analyses for female-biased (left panel **C**) and male-biased genes (right panel **C**). Gene enrichment analysis for female-biased genes based on the MSigDB Hallmark database (**D**). A volcano plot is shown for the differential protein abundance analysis between fibrous plaques from female and male patients (**E**). GO enrichment analyses for female-biased (left panel **F**) and male-biased proteins (right panel **F**). Boxplots representing key female-biased (**G**) and male-biased proteins (**H**).

To elucidate the specific transcriptomic differences between male and female plaques, we performed sex differential gene expression analyses on 191 fibrous and 187 atheromatous plaques (Supplemental Fig. 2D & E). A total of 20,902 protein-coding genes were measured in 61 female and 130 male fibrous plaques and 29 and 158 atheromatous plaques, respectively. Differential expression analysis in the fibrous plaques identified 138 differentially expressed genes between the sexes. Of these, 32 were higher expressed in female and 106 in male fibrous plaques (Fig. 3B, Supplemental Table 4 & 5). Genes higher expressed in female fibrous plaques were enriched for extracellular matrix (ECM) and collagen fibril organization, SMC migration, and nitric oxide metabolic process (all p<0.001, Fig. 3C, Supplemental Table 7). Additionally, there were strong signals towards endothelial-to-mesenchymal transition (EndMT), which was the top enrichment based on the MSigDB Hallmark database (p<0.001, Fig. 3D). The male fibrous genes were enriched for activation of immune response, leukocyte- and lymphocyte-mediated immunity, and antigen presentation (all p<0.001, Fig. 3C, Supplemental Table 7).

### Untargeted plaque proteomics further highlight EndMT as a key process in female fibrous plaques

To understand the importance of ECM organization in fibrous plaques from women compared to men, we generated untargeted proteomics data enriched for ECM proteins using LC-MS (see methods for details). Protein analysis was performed on 22 female and 44 male fibrous plaques and 9 female and 53 male atheromatous plaques (Supplemental Fig. 2F & G). Differential abundance analysis in the fibrous plaques identified 343 differentially expressed proteins, of which 211 were higher expressed in female and 132 in male fibrous plaques (Fig. 3E, Supplemental Table 4 & 9). Enrichment analyses linked female-biased proteins to cytoskeleton organization, actin filament-based movement, and muscle structure development (all p<0.001, Fig. 3F, Supplemental Table 11). These are key processes involved in EndMT. Proteins more abundant in fibrous plaques from women include MYH10, ACTN4, ENG, and TGFβ1 (Fig. 3G). Actin alpha 4 (ACTN4) primarily functions in cytoskeletal organization and cellular junctions, structural changes important for cell detachment and motility ^39^. Similar to ACTN4, myosin heavy chain (MYH10) is involved in cellular contractility and motility ^40^. Endoglin (ENG) is a co-receptor that enhances transforming growth factor beta 1 (TGFβ1) signaling, one of the most important stimuli for EndMT ^16,41^. The male-biased proteins were enriched for immune response, leukocyte- and lymphocyte-mediated immunity, and catabolic processes (all p<0.001, Fig. 3F, Supplemental Table 11). Proteins more abundant in fibrous plaques from men include CCL18, ICAM1, CXCL16, and MMP2. Chemokine (C-C motif) ligand 18 (CCL18) is a chemokine that contributes to the recruitment of immune cells to the site of plaque formation and has been linked to plaque instability through macrophages ^42^. Intercellular adhesion molecule 1 (ICAM1) facilitates the adhesion of leukocytes to ECs, promoting their migration into the vascular wall ^43,44^. C-X-C motif chemokine ligand 16 (CXCL16) is a chemokine that mediates the recruitment of these leukocytes to the plaque site and is involved in the uptake of oxLDL by macrophages ^45^. Finally, matrix metallopeptidase 2 (MMP2) is involved in the breakdown of ECM and is associated with unstable CAD ^46^.

### scRNAseq expression of sex-biased genes point to SMCs, ECs, and Macrophages

To investigate the contribution of different cell types to the observed gene expression and protein abundance differences between sexes, we used single-cell RNA sequencing (scRNAseq) data from carotid plaques of a separate subset of 46 patients from the same cohort (20 women and 26 men, Fig. 4A). Genes upregulated in female fibrous plaques were highest expressed in SMCs, while also showing high expression in endothelial cells (ECs) (Fig. 4B). In contrast, genes upregulated in male fibrous plaques showed highest expression in resident macrophages and inflammatory macrophages. scRNAseq data mapped the female fibrous proteins to SMC and endothelial cell phenotypes. In contrast, the male fibrous proteins were highest expressed in foam cells, resident macrophages, and inflammatory macrophages (Fig. 4B). In general, the cell type-specific expression from plaque proteomics closely mimicked those found in plaque transcriptomics, which was also seen for the female genes and proteins in the atheromatous comparison (Supplemental Fig. 3A & B).

**Figure 4.**
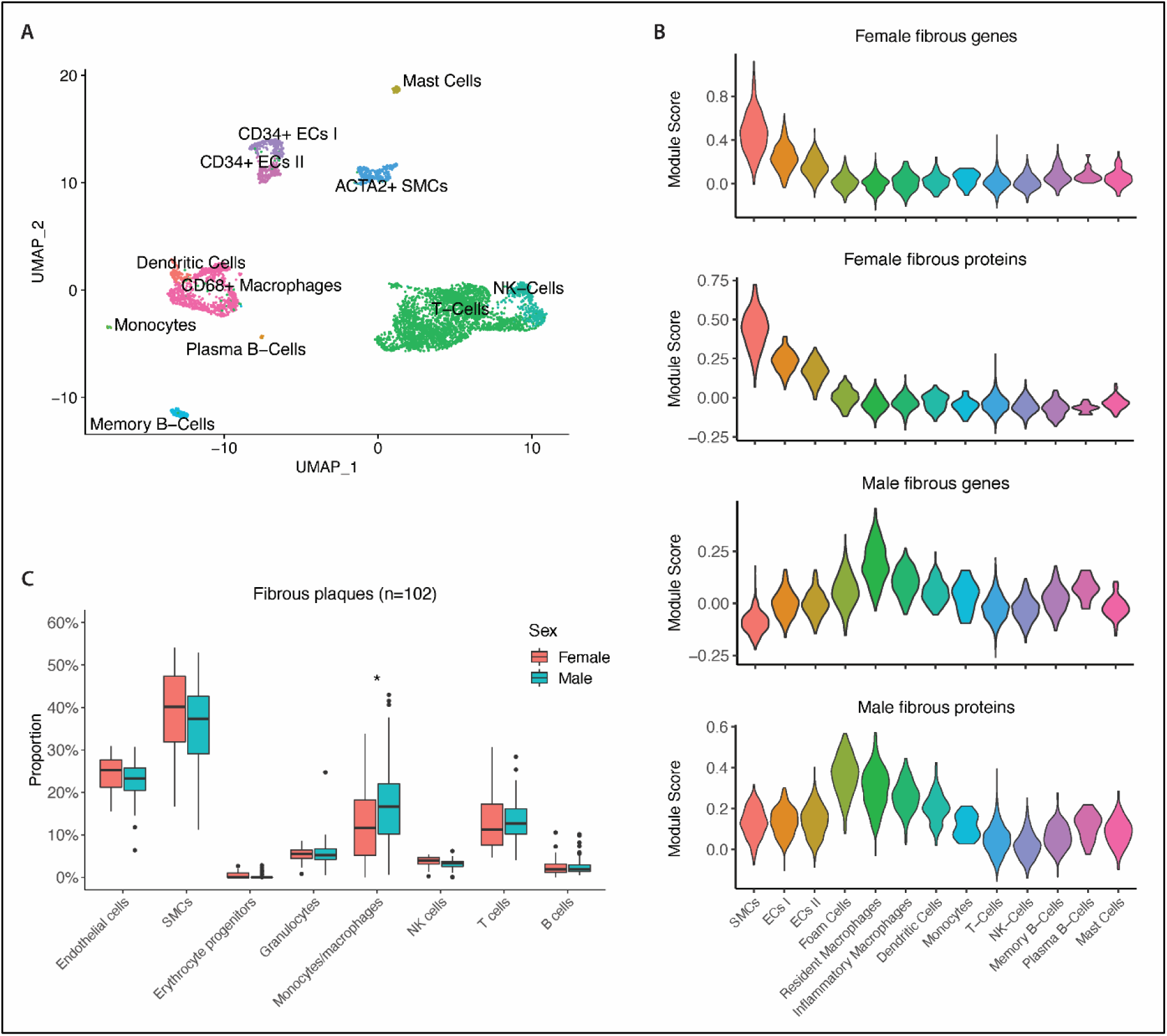
UMAP plot of 4,948 single cells from carotid plaques (20 female and 26 male patients) (**A**). Module score (see Methods) expression of female fibrous genes and proteins (upper two panels **B**) and male fibrous genes and proteins in plaque cell types (lower two panels **B**). Boxplot is shown for the sex-stratified deconvolution analysis on fibrous plaques using DNA methylation data. Proportion represents the predicted contribution (%) of each cell type to the fibrous plaque (**C**).

### Cell-type deconvolution based on DNA methylation supports a different ratio of immune cells/SMC+ECs between male and female fibrous plaques

To study if sex differences in cell-type-specific expression can be linked to differences in cellular composition, we performed deconvolution analyses on plaque DNA methylation data. A comprehensive DNA methylation atlas comprising 39 cell types sorted from 205 healthy tissue samples^47^, underwent filtering to select cells expected to be present in atherosclerotic plaques. This condensed methylation atlas, consisting of 8 selected cell types, served as a reference to estimate the cellular composition of 102 fibrous (Fig. 4C) and 117 atheromatous plaques (Supplemental Fig. 3C). These results indicate that fibrous cellular landscape mainly consists of SMCs (37%), ECs (23%), macrophages/monocytes (16%), T cells (13%) and granulocytes (6%). Male fibrous plaques contained significantly more macrophages and monocytes compared to their female counterparts. Although not significant, there appears to be a trend towards higher proportions of SMCs and ECs in female plaques. SMCs and macrophages emerged as primary drivers for inherent sex disparities in plaque transcriptomics and proteomics.

### Female fibrous plaques are characterized by endothelial-to-mesenchymal transition and associated to smoking through methylation changes

Cell transition within atherosclerotic plaques has been described as a contributing factor to plaque progression, especially in female plaques ^14–17^. To further understand how these sex differences arise, we used a gene expression signature that identifies EndMT single-cell lineages within human populations of plaque ECs and SMCs ^38^ (Fig. 5A). The expression of the female-biased fibrous genes along the trajectory of these populations revealed increasing expression patterns within the EndMT lineages (Fig. 5B). These lineages allowed us to pinpoint intermediate EndMT markers that are representative of the active ongoing transition, particularly those most expressed in the EC1 and SMC2 subtypes. EC1 cells are positive for the endothelial marker CD34 and the SMC marker *ACTA2*, with differential genes enriched for TGFβ response, SMC proliferation, and EndMT ^14^. Moreover, SMC2 cells express markers indicative of cellular migration, with differential genes enriched for TGFβ and BMP signalling. Notably, nearly half of the female fibrous genes (15 out of 32) overlapped with the identified intermediate markers for mid-stage EndMT ^38^ (Fig. 5C), showing increased expression in EC1 and SMC2 cells (Supplemental Fig. 3D).

**Figure 5.**
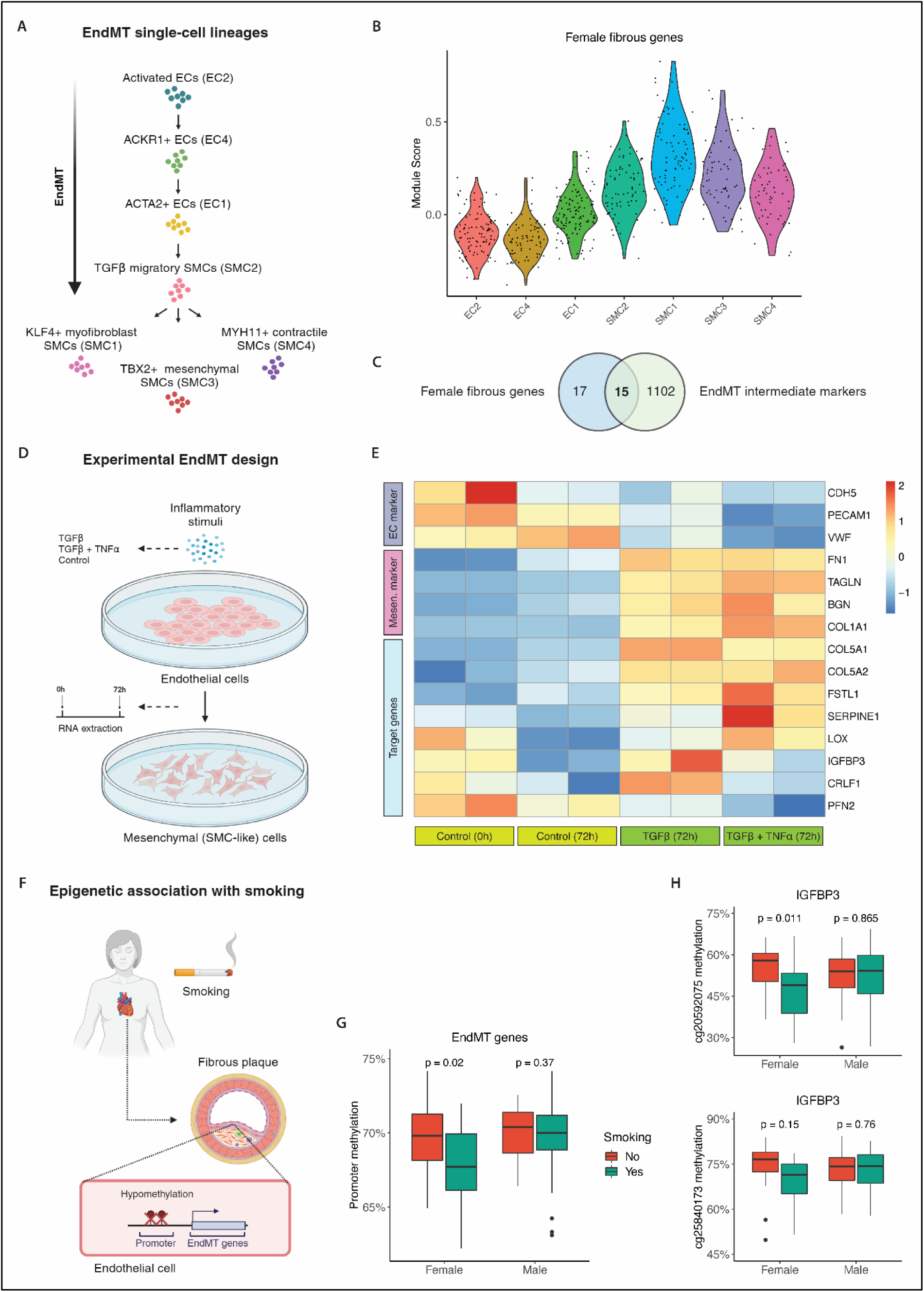
Identification of EndMT single-cell lineages in plaque ECs and SMCs using gene expression signatures^38^ (**A**). Violin plot showing the expression of female fibrous genes along the trajectory of EndMT lineages (**B**). Experimental design of EndMT in vitro model (see Methods): human coronary ECs were stimulated with inflammatory stimuli to induce transdifferentiation towards mesenchymal (SMC-like) cells (**C**). Heatmap displaying changes in the expression of known endothelial and mesenchymal markers, alongside our target genes (y-axis), across different conditions (x-axis) (**D**). Overview of the potential relation between smoking and EndMT promoter methylation changes in fibrous plaques from women (**E**). Boxplot representing the promoter methylation changes of EndMT genes in fibrous plaques (**F**). Boxplots representing the promoter methylation changes of EndMT gene IGFBP3 in fibrous plaques (**G**).

To further investigate how these genes relate to EndMT, we experimentally studied the expression of these 15 target genes, along with known EC and mesenchymal markers in an in vitro model of EndMT. Human coronary endothelial cells were stimulated to undergo EndMT under two conditions: TGFβ alone, and a combination of TGFβ with TNFα (Fig. 5D). The results demonstrated clear differential gene expression responses (Fig. 5E). All endothelial markers decreased in expression under stimulation with TGFβ and TNFα (mean logFC: -2.4, P-adj<0.001), whereas all mesenchymal markers increased (mean logFC: 2.3, P-adj<0.001). Similar expression patterns were observed with just TGFβ stimulation. Out of the seven target genes that had a minimal average expression, we identified three genes that had a positive effect size under both stimulating conditions (mean logFC: 0.5), of which *COL5A1* and *FSTL1* were both found to be significantly increased (P-adj<0.05).

Smoking is associated with fibrous plaques in women (Fig. 1C) and can have strong effects on DNA methylation in plaque tissue (Fig. 5F)^13^. To investigate the effect of smoking on the regulation of our 15 target genes, we obtained DNA methylation data from 38 female and 64 male fibrous plaques. We specifically examined CpGs in the promoter regions (TSS ≤ 1kb) of the 15 EndMT genes, resulting in 9 promoter CpGs associated with 4 genes (3 CpGs with *COL5A1*, 2 CpGs with *IGFBP3*, 2 CpGs with *PCOLCE2* and 2 CpGs with *FSTL1*). We found that these promoter regions are significantly hypomethylated in women who smoke and have fibrous plaques compared to females with fibrous plaques that do not smoke (p=0.02) (Fig. 5G). Upon closer examination by gene, the promoter region of *IGFBP3* was significantly hypomethylated (cg20592075 & cg25840173) (Fig. 5H). *IGFBP3* is a known GWAS locus gene for coronary artery calcification (CAC)^48^ and a mesenchymal marker indicative of EndMT. This hypomethylation was not observed in male plaques, suggesting that the observed differences in the molecular drivers of fibrous plaques may be explained by sex specific differences in response to risk factors.

## Discussion

In this study, we show that patients presenting with late-stage atherosclerotic plaques, histologically scored as fibrous, exhibit sex differences in risk factors. Women are more likely to be smokers with higher LDL and CRP plasma levels, while men are more likely to have diabetes and history of CAD. Our data show that female fibrous plaques are characterized by EndMT and SMC-driven ECM remodelling, whereas male fibrous plaques point to inflammation driven by macrophages. We experimentally validated female-biased EndMT gene signatures in vitro and linked them to smoking-mediated promoter methylation changes, suggesting a sex-dependent epigenetic response to risk factors.

While there has been a general decline in tobacco use in recent decades, this reduction has been less significant for women compared to men ^49^. Notably, women who smoke have a 25% increased risk of developing CAD compared to men^12^, and prolonged smoking, even at lower levels, results in worse cardiovascular outcome ^11,50^. Although the mechanisms underlying these differences remain unclear, previous studies have demonstrated that smoking is associated with plaque erosion in women, which might explain the increased risk of ACSs ^1^. In this study, we found evidence that women with fibrous plaque are more likely to smoke compared to men with fibrous plaques (41% vs. 33%). While sex differences in the prevalence of plaque erosion and fibrous plaques are especially pronounced at younger ages (<50 years)^1,51^, we show that women in our cohort (average age 68) also tend to have fibrous plaques more frequently than men (50% vs. 32%), as consistently reported over the years ^8,22^. Men with fibrous plaques, on the other hand, are more likely to have diabetes (29% vs. 20%). Patients with diabetes were shown to have a similar prevalence of plaque erosion and rupture as patients without diabetes while at the same time having more atheromatous-like plaque features, such as the accumulation of macrophages and lipids ^52^.

While histology has long been and continues to be pivotal in identifying structural differences in atherosclerotic plaques, including those related to sex, the integration of advanced sequencing technologies and multi-omics analyses has revealed a deeper molecular complexity. Recent sex-stratified studies indicate that male and female plaques harbour distinct molecular and cellular mechanisms and underscore the complex interplay with hormone dynamics in shaping sex-specific atherosclerosis beyond the surface level of histological features ^14,15^. Consistent with this theory, our multi-omics study demonstrates that patients presenting with late-stage fibrous atherosclerotic plaques, despite being histologically similar, reveal distinct biological processes when stratifying by sex. We find that these differences are linked between multiple biological layers, from RNA to ECM proteins, and from the impact of smoking on epigenetic changes to gene expression. A recent study investigating the effect of smoking on the plaque transcriptome revealed a sex-dependent upregulation of SMC gene *CRLF1* in women, which is involved in ECM remodelling and exhibits the highest expression in fibrous-like plaque types ^53^. In-depth analyses of fibrous plaques at protein, gene, and DNA methylation levels revealed female fibrous plaques characterized by EndMT and SMC-driven ECM remodelling. In contrast, male fibrous plaques point to inflammation driven by macrophages. This aligns with previously identified sex differences in SMCs, ECs, and macrophages at the molecular level ^14,16^, indicating that deeper analyses point towards diverging pathological mechanisms between the sexes even without clear histological differences. This observation aligns with the distribution of molecular-based plaque phenotypes within our study population, where atheromatous-like phenotypes (lipomatous and fibro-inflammatory) were more prevalent in fibrous plaques from men than women. It has been previously described that intrinsic differences in molecular mechanisms might influence the choice of drug treatments, particularly based on the molecular profiling for tumor subtypes in cancer ^54^. Similarly, the differences in mechanisms found in this study may have consequences for the domain of applications in emerging antifibrotic or anti-inflammatory therapies ^55,56^, which may vary in efficacy by sex and plaque phenotype.

EndMT has been previously linked to atherosclerosis processes ^16,17^. Several of the analyses in this study highlight EndMT as a leading process involved in end-stage fibrous plaques in women, where an enrichment of EndMT-associated genes and proteins was observed. scRNAseq and methylation deconvolution further reveal a more prominent role of ECs and SMCs in fibrous plaques in women compared to men. Recently published intermediate human EndMT markers ^38^ were enriched within the female-biased fibrous genes in this study, also showing increased expression in transitioning cells, reinforcing the main enrichment of EndMT that was found in fibrous plaques from women. Finally, we experimentally validated the involvement of these genes in EndMT in human coronary endothelial cells. All of these results combined suggest a potential role of EndMT in driving sex differences in fibrous atherosclerosis. As endothelial cells lose integrity and detach during EndMT, one might envision a role in the pathology that leads to plaque erosion ^57^. In both EndMT and plaque erosion, endothelial activation is triggered by disturbed flow, endothelial shear stress, oxidative stress, and inflammatory stimuli ^5,17^. Although the suggestions have been made before ^58^, more research is required to investigate the exact relationship between EndMT and the disrupted endothelial layer that results in plaque erosion.

Using our plaque epigenetics data, we provide evidence that the average methylation of EndMT gene promoters involved in fibrous female plaques, is significantly lower in fibrous plaques from women who smoke. Notably, this hypomethylation was not observed in male counterparts, regardless of their smoking status. Moreover, these findings align with previous work on plaque epigenetics, underscoring how smoking can significantly impact genome-wide DNA methylation^13^ and gene expression^53^, and influence epigenetic regulation in atherosclerotic disease. Furthermore, it has previously been demonstrated that the expression of EndMT genes is mediated through promoter hypomethylation upon TGF-β stimulation, a potent inducer of EndMT ^59^. Although the exact relationship between risk factors, epigenetics, and atherosclerotic pathways requires further investigation, our findings hint towards a sex-dependent epigenetic response to smoking that might be involved in EndMT.

This study has limitations. Firstly, the histological plaque sections were scored using semi-quantitative methods. This may be considered a limitation, given its relatively low resolution compared to the deeper molecular complexity at protein, gene, and DNA methylation levels explored in this study. However, these histopathological measures are widely used for plaque phenotyping, and demonstrate good replication metrics and intra-and interobserver reliability ^18,19,22,23^. Additionally, the classification of plaques into fibrous and atheromatous phenotypes was primarily based on the presence and size of the lipid core. However, other histopathological characteristics have also been recognized as useful for classification. To address this, we incorporated the plaque vulnerability index, a combined score of individual histological features including SMC content, collagen content, lipid content, macrophage content, and intraplaque hemorrhage ^14^. Notably, this index revealed substantial differences between fibrous and atheromatous plaques (2.3 vs. 4.3, p<0.001 ) yet showed no sex differences within these groups (Table 1, Supplemental Table 1), thereby supporting our classification method. Another limitation is the underrepresentation of women in the Athero-Express biobank, where only one in three patients is a woman. Similarly, small numbers of women were observed in the protein (24%), bulk RNA (24%), single-cell RNA (39%), and DNA methylation data (27%), which decreases the statistical power of these analyses. However, the signals identified were statistically robust and consistent across biological layers. We also acknowledge that the plaques analyzed are isolated at the final stage of atherosclerosis, representing the culmination of disease progression over time.

Consequently, while we identify EndMT as a predominant process in fibrous plaques of women, it remains unclear at which stage of plaque development this process becomes significant. Moreover, it is important to note that the methylation observed at the EndMT promoters reflects a composite of signals from all cell types present in the plaque. Finally, our results may be influenced by the specific segment locations within each plaque. Different regions within a plaque can exhibit different cellular contributions, potentially explaining distinct molecular mechanisms. To minimize this variability, only plaque segments adjacent to the culprit lesion were used for sequencing, and this protocol was followed in both males and female plaques.

In conclusion, patients with end-stage fibrous atherosclerotic plaques reveal distinct sex-specific clinical and molecular profiles. Female plaques are characterized by SMC-driven ECM remodeling and EndMT linked to smoking-induced epigenetic changes, whereas male plaques are driven by macrophage-driven inflammation, associating with a higher prevalence of diabetes. These mechanisms could be key to understanding plaque erosion at the molecular level and offer novel targets for atherosclerosis therapies, as they account for both sex and plaque phenotype.

## Data Availability

Data will be made available upon reasonable request.

## Acknowledgements

The authors would like to acknowledge all the participants of the Athero-Express biobank for agreeing to be part of the study.

## References

1. Sato, Y. et al. Sex Differences in Coronary Atherosclerosis. Curr Atheroscler Rep 24, 23–32 (2022).

2. Libby, P., Pasterkamp, G., Crea, F. & Jang, I.-K. Reassessing the Mechanisms of Acute Coronary Syndromes. Circ Res 124, 150–160 (2019).

3. van Lammeren, G. W. et al. Time-Dependent Changes in Atherosclerotic Plaque Composition in Patients Undergoing Carotid Surgery. Circulation 129, 2269–2276 (2014).

4. Farb, A. et al. Coronary Plaque Erosion Without Rupture Into a Lipid Core. Circulation 93, 1354–1363 (1996).

5. Fahed, A. C. & Jang, I.-K. Plaque erosion and acute coronary syndromes: phenotype, molecular characteristics and future directions. Nat Rev Cardiol 18, 724–734 (2021).

6. Quillard, T., Franck, G., Mawson, T., Folco, E. & Libby, P. Mechanisms of erosion of atherosclerotic plaques. Curr Opin Lipidol 28, 434–441 (2017).

7. Kolte, D., Libby, P. & Jang, I.-K. New Insights Into Plaque Erosion as a Mechanism of Acute Coronary Syndromes. JAMA 325, 1043 (2021).

8. Hellings, W. E. et al. Gender-associated differences in plaque phenotype of patients undergoing carotid endarterectomy. J Vasc Surg 45, 289–296 (2007).

9. Sakkers, T. R. et al. Sex differences in the genetic and molecular mechanisms of coronary artery disease. Atherosclerosis 384, 117279 (2023).

10. Low Wang, C. C., Hess, C. N., Hiatt, W. R. & Goldfine, A. B. Clinical Update: Cardiovascular Disease in Diabetes Mellitus. Circulation 133, 2459–2502 (2016).

11. Appelman, Y., van Rijn, B. B., ten Haaf, M. E., Boersma, E. & Peters, S. A. E. Sex differences in cardiovascular risk factors and disease prevention. Atherosclerosis 241, 211–218 (2015).

12. Huxley, R. R. & Woodward, M. Cigarette smoking as a risk factor for coronary heart disease in women compared with men: a systematic review and meta-analysis of prospective cohort studies. The Lancet 378, 1297–1305 (2011).

13. Siemelink, M. A., et al. Smoking is Associated to DNA Methylation in Atherosclerotic Carotid Lesions. Circ Genom Precis Med 11, (2018).

14. Diez Benavente, Ernest Karnewar, S., et al. Female gene networks are expressed in myofibroblast-like smooth muscle cells in vulnerable atherosclerotic plaques. BioRxiv (2023).

15. Hartman, R. J. G. et al. Sex-Stratified Gene Regulatory Networks Reveal Female Key Driver Genes of Atherosclerosis Involved in Smooth Muscle Cell Phenotype Switching. Circulation 143, 713–726 (2021).

16. Wesseling, M., Sakkers, T. R., de Jager, S. C. A., Pasterkamp, G. & Goumans, M. J. The morphological and molecular mechanisms of epithelial/endothelial-to-mesenchymal transition and its involvement in atherosclerosis. Vascul Pharmacol 106, 1–8 (2018).

17. Evrard, S. M. et al. Endothelial to mesenchymal transition is common in atherosclerotic lesions and is associated with plaque instability. Nat Commun 7, 11853 (2016).

18. Verhoeven, B. A. N. et al. Athero-express: Differential atherosclerotic plaque expression of mRNA and protein in relation to cardiovascular events and patient characteristics. Rationale and design. Eur J Epidemiol 19, 1127–1133 (2004).

19. van Koeverden, I. D. et al. Testosterone to oestradiol ratio reflects systemic and plaque inflammation and predicts future cardiovascular events in men with severe atherosclerosis. Cardiovasc Res 115, 453–462 (2019).

20. van der Laan, S. W. et al. Variants in ALOX5, ALOX5AP and LTA4H are not associated with atherosclerotic plaque phenotypes: The Athero-Express Genomics Study. Atherosclerosis 239, 528–538 (2015).

21. Idänpään-Heikkilä, J. E. Ethical principles for the guidance of physicians in medical research--the Declaration of Helsinki. Bull World Health Organ 79, 279 (2001).

22. de Bakker, M. et al. The age- and sex-specific composition of atherosclerotic plaques in vascular surgery patients. Atherosclerosis 310, 1–10 (2020).

23. Hellings, W. E. et al. Intraobserver and interobserver variability and spatial differences in histologic examination of carotid endarterectomy specimens. J Vasc Surg 46, 1147– 1154 (2007).

24. Mokry, M. et al. Transcriptomic-based clustering of human atherosclerotic plaques identifies subgroups with different underlying biology and clinical presentation. Nature Cardiovascular Research 1, 1140–1155 (2022).

25. Love, M. I., Huber, W. & Anders, S. Moderated estimation of fold change and dispersion for RNA-seq data with DESeq2. Genome Biol 15, 550 (2014).

26. Yu, G., Wang, L.-G., Han, Y. & He, Q.-Y. clusterProfiler: an R Package for Comparing Biological Themes Among Gene Clusters. OMICS 16, 284–287 (2012).

27. Naba, A. et al. The extracellular matrix: Tools and insights for the “omics” era. Matrix Biology 49, 10–24 (2016).

28. Ritchie, M. E. et al. limma powers differential expression analyses for RNA-sequencing and microarray studies. Nucleic Acids Res 43, e47–e47 (2015).

29. Depuydt, M. A. C. et al. Microanatomy of the Human Atherosclerotic Plaque by Single-Cell Transcriptomics. Circ Res 127, 1437–1455 (2020).

30. Slenders, L. et al. Intersecting single-cell transcriptomics and genome-wide association studies identifies crucial cell populations and candidate genes for atherosclerosis. European Heart Journal Open 2, (2022).

31. Stuart, T. & Satija, R. Integrative single-cell analysis. Nat Rev Genet 20, 257–272 (2019).

32. Chen, E. Y. et al. Enrichr: interactive and collaborative HTML5 gene list enrichment analysis tool. BMC Bioinformatics 14, 128 (2013).

33. Diez Benavente, E., et al. Atherosclerotic Plaque Epigenetic Age Acceleration Predicts a Poor Prognosis and Is Associated With Endothelial-to-Mesenchymal Transition in Humans. Arterioscler Thromb Vasc Biol 44, 1419–1431 (2024).

34. Maksimovic, J., Phipson, B. & Oshlack, A. A cross-package Bioconductor workflow for analysing methylation array data. F1000Res 5, 1281 (2017).

35. Moss, J. et al. Comprehensive human cell-type methylation atlas reveals origins of circulating cell-free DNA in health and disease. Nat Commun 9, 5068 (2018).

36. Zeisberg, E. M. et al. Endothelial-to-mesenchymal transition contributes to cardiac fibrosis. Nat Med 13, 952–961 (2007).

37. Yoshimatsu, Y. et al. TNF-α enhances TGF-β-induced endothelial-to-mesenchymal transition via TGF-β signal augmentation. Cancer Sci 111, 2385–2399 (2020).

38. Slenders L, et al. Identification of endothelial-to-mesenchymal transition gene signatures in single-cell transcriptomics of human atherosclerotic tissue. bioRxiv (2023).

39. Tentler, D., Lomert, E., Novitskaya, K. & Barlev, N. A. Role of ACTN4 in Tumorigenesis, Metastasis, and EMT. Cells 8, 1427 (2019).

40. Wang, Y. et al. Myosin Heavy Chain 10 (MYH10) Gene Silencing Reduces Cell Migration and Invasion in the Glioma Cell Lines U251, T98G, and SHG44 by Inhibiting the Wnt/β-Catenin Pathway. Medical Science Monitor 24, 9110–9119 (2018).

41. Chen, P.-Y. et al. Endothelial TGF-β signalling drives vascular inflammation and atherosclerosis. Nat Metab 1, 912–926 (2019).

42. Singh, A. et al. CCL18 aggravates atherosclerosis by inducing CCR6-dependent T-cell influx and polarization. Front Immunol 15, (2024).

43. Poston, R. N., Haskard, D. O., Coucher, J. R., Gall, N. P. & Johnson-Tidey, R. R. Expression of intercellular adhesion molecule-1 in atherosclerotic plaques. Am J Pathol 140, 665–73 (1992).

44. Alvandi, Z. & Bischoff, J. Endothelial-Mesenchymal Transition in Cardiovascular Disease. Arterioscler Thromb Vasc Biol 41, 2357–2369 (2021).

45. Korbecki, J. et al. The Role of CXCL16 in the Pathogenesis of Cancer and Other Diseases. Int J Mol Sci 22, 3490 (2021).

46. Samah, N., Ugusman, A., Hamid, A. A., Sulaiman, N. & Aminuddin, A. Role of Matrix Metalloproteinase-2 in the Development of Atherosclerosis among Patients with Coronary Artery Disease. Mediators Inflamm 2023, 1–16 (2023).

47. Loyfer, N. et al. A DNA methylation atlas of normal human cell types. Nature 613, 355–364 (2023).

48. Kavousi, M. et al. Multi-ancestry genome-wide study identifies effector genes and druggable pathways for coronary artery calcification. Nat Genet 55, 1651–1664 (2023).

49. Garcia, M., Mulvagh, S. L., Bairey Merz, C. N., Buring, J. E. & Manson, J. E. Cardiovascular Disease in Women. Circ Res 118, 1273–1293 (2016).

50. Peters, S. A. E., Carcel, C., Millett, E. R. C. & Woodward, M. Sex differences in the association between major risk factors and the risk of stroke in the UK Biobank cohort study. Neurology 95, (2020).

51. Yahagi, K., Davis, H. R., Arbustini, E. & Virmani, R. Sex differences in coronary artery disease: Pathological observations. Atherosclerosis 239, 260–267 (2015).

52. Sugiyama, T. et al. Coronary Plaque Characteristics in Patients With Diabetes Mellitus Who Presented With Acute Coronary Syndromes. J Am Heart Assoc 7, (2018).

53. Lan, T. et al. Tobacco smoking is associated with sex- and plaque-type specific upregulation of CRLF1 in atherosclerotic lesions. Atherosclerosis 397, 118554 (2024).

54. Malone, E. R., Oliva, M., Sabatini, P. J. B., Stockley, T. L. & Siu, L. L. Molecular profiling for precision cancer therapies. Genome Med 12, 8 (2020).

55. Ridker, P. M. et al. Antiinflammatory Therapy with Canakinumab for Atherosclerotic Disease. New England Journal of Medicine 377, 1119–1131 (2017).

56. Nidorf, S. M. et al. Colchicine in Patients with Chronic Coronary Disease. New England Journal of Medicine 383, 1838–1847 (2020).

57. Tombor, L. S. et al. Single cell sequencing reveals endothelial plasticity with transient mesenchymal activation after myocardial infarction. Nat Commun 12, 681 (2021).

58. Kovacic, J. C. et al. Endothelial to Mesenchymal Transition in Cardiovascular Disease: JACC State-of-the-Art Review. J Am Coll Cardiol 73, 190–209 (2019).

59. Cardenas, H. et al. TGF-β induces global changes in DNA methylation during the epithelial-to-mesenchymal transition in ovarian cancer cells. Epigenetics 9, 1461–1472 (2014).

